# Biomarkers and Outcomes in Hospitalized Covid-19 Patients: A Prospective Registry

**DOI:** 10.1101/2022.07.20.22277718

**Authors:** Raghubir S Khedar, Rajeev Gupta, Krishna Kumar Sharma, Kartik Mittal, Harshad C Ambaliya, Jugal B Gupta, Surendra Singh, Swati Sharma, Yogendra Singh, Alok Mathur

## Abstract

**Objectives:** To determine association of biomarkers high sensitivity C-reactive protein (hsCRP), D-dimer, interleukin-6 (IL-6), lactic dehydrogenase (LDH), ferritin and neutrophil-lymphocyte ratio (NLR) at hospital admission with clinical features and outcomes in Covid-19.

**Methods:** Successive virologically confirmed Covid-19 patients hospitalized from April 2020 to July 2021 were recruited in a prospective registry. Details of clinical presentation, investigations, management and outcomes were recorded. All the biomarkers were divided into tertiles to determine associations with clinical features and outcomes. Numerical data are presented in median and interquartile range (IQR 25-75). Univariate and multivariate (age, sex, risk factor, comorbidity adjusted) odds ratio (OR) and 95% confidence intervals (CI) were calculated to determine association of deaths with each biomarker.

**Results:** We identified 3036 virologically confirmed Covid-19 patients during the study period, 1215 were hospitalized and included in the present study. Men were 70.0%, aged >60y 44.8%, hypertension 44.8% diabetes 39.6% and cardiovascular disease 18.9%. Median symptom duration was 5 days (IQR 4-7) and SpO_2_ 95% (90-97). Total white cell count was 6.9×10^3^/µl, (5.0-9.8), neutrophils 79.2% (68.1-88.2) and lymphocytes 15.8% (8.7-25.5). Medians (IQR) for biomarkers were hsCRP 6.9 mg/dl (2.2-18.9), D-dimer 464 ng/dl (201-982), IL-6 20.1 ng/dl (6.5-60.4), LDH 284 mg/dl (220-396) and ferritin 351 mg/dl (159-676). Oxygen support at admission was in 38.6%, and non-invasive or invasive ventilatory support in 11.0% and 11.6% respectively. 173 (13.9%) patients died and 15 (1.2%) transferred to hospice care. For each biomarker, those in the second and third tertiles, compared to the first, had worse clinical and laboratory abnormalities, and greater oxygen and ventilatory support. Multivariate adjusted OR (95% CI) for deaths in second and third vs first tertiles, respectively, were for hsCRP 2.29(1.14-4.60) and 13.39(7.23-24.80); D-dimer 3.26(1.31-7.05) and 13.89(6.87-28.27); IL-6 2.61(1.31-5.18) and 10.96(5.88-20.43); ferritin 3.19(1.66-6.11) and 9.13(4.97-16.78); LDH 1.85(0.87-3.97) and 10.51(5.41-20.41); and NLR 3.34(1.62-6.89) and 17.52(9.03-34.00) (p<0.001).

**Conclusions:** In Covid-19, high levels of biomarkers-hsCRP, D-dimer, IL-6, LDH, ferritin and NLR are associated with more severe illness and significantly greater in-hospital mortality. NLR, a simple, widely available and inexpensive investigation provides prognostic information similar to the more expensive biomarkers.

## INTRODUCTION

The pandemic of severe acute coronary syndrome coronavirus-2 (SARS-CoV-2) infection and Covid-19 continues unabated in many regions of the world.^1^ The current wave of the epidemic has been triggered by the omicron-variant of Covid-19 including BA.1, BA.2, BA.1.12 BA.2.75, BA.4 and BA.5 although previous variants (ancestral, alpha, beta, gamma and delta) are also present.^2,3^ Although adverse outcomes in the present omicron-wave is not as severe as the delta-variant, a significant number of patients are being hospitalized and high transmission of these variants has resulted in more hospitalizations as compared to the previous waves in some countries.^3-6^

A number of prognostic markers have been identified to indicate severity of Covid-19.^7,8^ These include clinical markers such as tachypnea, tachycardia and hypoxia, radiological abnormalities, and laboratory-based biomarkers. Biomarkers include interleukins (IL) 6, 4 and 10, procalcitonin, C-reactive protein (CRP), serum amyloid A (SAA), neutrophil, lymphocyte, monocyte and platelet counts, alanine aminotransferase (ALT), aspartate aminotransferase (AST), lactic dehydrogenase (LDH), creatinine kinase (CK), CK-MB isoenzyme, activated partial thromboplastin time (aPTT) and prothrombin time.^7^ Thrombotic biomarkers are also important and are markers of platelet activation, platelet aggregation, endothelial cell activation or injury, coagulation and fibrinolysis and include fibrinogen, D-dimer and aPTT.^8^ A systematic review and meta-analysis reported significant association of lymphopenia, thrombocytopenia and elevated levels of CRP, procalcitonin, LDH and D-dimer with adverse outcomes in Covid-19.^9^

Limited studies have evaluated the prognostic biomarkers in developing countries where the burden of Covid-19 is the highest.^10^ Only small studies are available from developing countries including India,^11-17^ where the disease led to massive burden and deaths. Measurement of laboratory biomarkers is expensive and not routinely available in most developing countries. We initiated a prospective Covid-19 registry at our hospital to evaluate disease pattern and outcomes.^18^ The present study aims to evaluate association of select biomarkers-high sensitivity C-reactive protein (hsCRP), D-dimer, IL-6, LDH, ferritin and neutrophil-lymphocyte ratio (NLR) at hospital admission with clinical presentation, investigations and outcomes in Covid-19. We also evaluated whether NLR, a low cost and widely available biomarker is as important as others for prognostication.

## METHODS

We initiated a registry of all patients with virologically confirmed Covid-19 admitted to our hospital since April 2020.^18^ The protocol was approved by the ethics committee of Eternal Heart Care Centre and Research Institute, Jaipur, India (Government of India registration, CDSCO No. ECR/615/Inst/RJ/2014/RR-20) before initiation of the study. Informed consent from all the patients or next-of-kin was obtained for anonymized data publication. The whole process was approved by the ethics committee.

### Setting

This is a 220-bed tertiary care hospital with major focus on critical care and cardiovascular sciences. It was designated an Advanced Covid Care Hospital by Government of Rajasthan and more than 20% beds in general wards and intensive care units were initially reserved for Covid-19 patients. This proportion was later increased to 50% and 75% as the number of critically-ill patients in the state increased. The hospital subsidized treatment to all the admitted patients according to the state government regulations. We developed a protocol for admission so that only those patients fulfilling definite clinical criteria were hospitalized. The case-report form was updated and modified from that available at the Regional Covid Care Hospital at local Health Sciences University.^19^

### Patients

All patients presenting to medical and emergency departments with symptoms suggestive of upper respiratory infections were screened with reverse transcriptase-polymerase chain reaction (RT-PCR). Details of the sample collection and testing protocol have been reported.^20^ We obtained details of case history, vital monitoring for all patients was recorded and a flow chart of all in-hospital investigations maintained. Hematological investigations included complete blood counts performed using XS-1000i, a 6-part fully automated analyzer from Sysmex Inc, USA. Biochemical investigations focused on measurement of blood glucose, renal function tests (urea, creatinine, electrolytes, uric acid) and liver function tests (bilirubin, AST, ALT, alkaline phosphatase, proteins, albumin, gamma glutaryl transferase) that were measured using Roche Cobas 6000, a fully automated molecular biochemistry and immunoassay analyzer. Covid-19 specific biomarkers measured at admission were hsCRP, IL-6, LDH and ferritin using Roche Cobas 6000. D-dimer was measured using a fully automated coagulation analyzer, ECL-760 ERBA and ACL Pro from Instrumentation Laboratories, USA.

All the hospitalized patients received clinical management according to national and international protocols.^18^ Essentially, hydration and oxygenation were maintained using oral or intravenous fluids and nasal-cannula based oxygen supplementation provided as needed. Steroids were used only in patients needing non-invasive or invasive ventilatory support. Remdesivir was used according to international guidelines.^21,22^ Anti-interleukin drugs were used infrequently and nonevidence-based therapies such as oral hydroxychloroquine, ivermectin, anti-viral drugs (e.g., favipiravir or ritonavir) or plasma therapy not recommended for hospitalized patients.

### Statistical analyses

All the data were computerized and entered into MS-Excel work-sheets. We focused on clinical history at presentation, admission hematological and biochemical investigations, radiological imaging-computerized tomographic (CT) scan, medical therapies, oxygenation, ventilation and in-hospital outcomes. Long-term follow-up data are not yet available. Descriptive analyses have been performed using SPSS package (Version 21.0). The categorical variables have been reported as numbers and percent while continuous variables are reported as medians and 25-75^th^ percentile interquartile range (IQR). As most of the biochemical variables had a skewed distribution, we used medians and IQR for descriptive statistics. Inter-groups comparisons have been performed using χ^2^-test for categorical variables and Kruskal-Wallis’ test for non-parametric continuous variables. The biochemical variables (hsCRP, IL-6, LDH, D-dimer, ferritin) and hematological variables (NLR) were divided into tertiles and clinical and other details tabulated accordingly. To determine odds ratios (OR) and 95% confidence intervals (95% CI) for deaths in second and third tertiles of various biomarkers vs first tertile patients, we initially performed a univariate logistic regression. Multivariate logistic regression was performed using variables likely to confound the outcomes such as age, sex, risk factors and comorbidities. We did not adjust for disease severity and other outcome measures, as these were the primary outcomes in the present study and likely to attenuate the OR’s and make comparisons of disease severity difficult. To determine inter-correlation of various biomarkers we developed a correlation matrix using Spearman’s test (rho). P values < 0.05 are considered significant.

## RESULTS

In the successive waves of Covid-19 epidemic from April 2020 to July 2021, we evaluated 16,146 suspected patients with nasopharyngeal samples for SARS-CoV-2 antigen RT-PCR test. Of these 3036 (18.8%) tested positive for the virus and 1251 (41.2%) were hospitalized. Data of these 1251 successively admitted patients have been obtained and analyzed. Men were 876 (70.0%), aged >60 years were 560 (44.8%) and prevalence of important comorbidities were in-hypertension 551 (44.1%), diabetes 495 (39.6%), cardiovascular disease 241 (18.9%), and asthma or chronic obstructive pulmonary disease (COPD) in 72 (2.7%). Median symptom duration was 5 days (IQR 4-7) and SpO_2_ at admission was 95% (90-97%). Total white cell count was 6.9×10^3^/µl (5.0-9.8), neutrophils 79.2% (68.1-88.2), lymphocytes 15.8% (8.7-25.5), and platelets 2.2×10^6^/µl (1.7-2.8). Levels of various biomarkers were CRP 6.9 mg/dl (2.2-18.9), IL-6 20.1 ng/dl (6.5-60.4), D-dimer 464 ng/dl (201-982), LDH 284 mg/dl (220-396) and ferritin 351 mg/dl (159-676). Details of renal and liver functions tests and radiography have been reported earlier.^17^ Oxygen support at admission was in 480 (38.6%), and during hospitalization nasal cannula based oxygen support was in 357 (28.7%), non-rebreather masks in 139 (11.2%), non-invasive ventilatory support in 137 (11.0%) and invasive ventilation in 144 (11.6%). Prone positioning (proning) was performed in 676 (54.3%) patients. Of the total hospitalized patients 1055 (84.9%) were discharged to home-based care, 15 (1.2%) transferred to hospice care and death was in 173 (13.9%).

The patients have been divided into tertiles of various biomarkers-hsCRP (Table 1, n=1130), D-dimer (Table 2, n=861), IL-6 (Table 3, n=1038), LDH (Table 4, n=721), ferritin (Table 5, n=998), and NLR (Table 6, n=1215). Details of demography, risk factors, comorbid conditions, clinical features and investigations, medical management and outcomes for each of the biomarker are provided in Tables 1-6. For each biomarker, those in the highest tertile have the most adverse clinical characteristics and significantly greater levels of clinical abnormalities-fever, shortness of breath, hypoxia, leukocytosis, lymphopenia- and higher levels of other biomarkers. Uses of steroids, anticoagulants, remdesivir and anti-inflammatory drugs as well as proning, oxygenation and non-invasive and invasive ventilatory support is also greater in those in the second and third tertiles (Table 1-6). There is a strong correlation among various biomarkers (Table 7). Results of parametric (Pearson’s) and non-parametric (Spearman’s) correlation among various biomarkers shows significant inter-correlation among all the biomarkers (p<0.001). Non-parametric analysis shows greater correlation (rho) of hsCRP with IL-6 (0.42), LDH (0.54), ferritin (0.42) and NLR (0.48) and NLR with hsCRP (0.48), LDH (0.48) and ferritin (0.38) (Table 7).

**Table 1:**
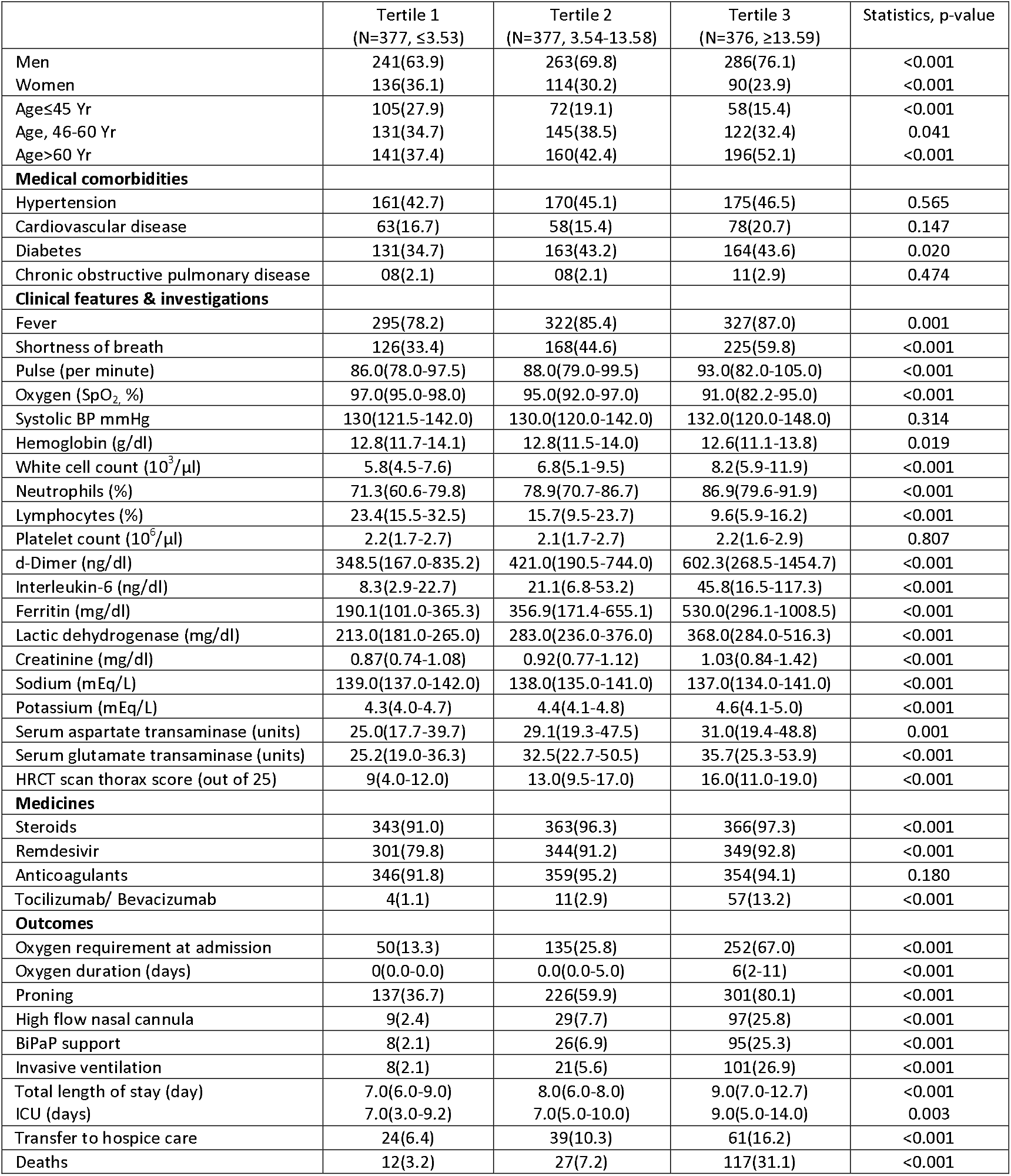
High sensitivity C-reactive protein (hsCRP) tertiles, clinical features and outcomes.

|  | Tertile 1<br>(N=377, ≤3.53) | Tertile 2<br>(N=377, 3.54-13.58) | Tertile 3<br>(N=376, ≥13.59) | Statistics, p-value |
| --- | --- | --- | --- | --- |
| Men | 241(63.9) | 263(69.8) | 286(76.1) | <0.001 |
| Women | 136(36.1) | 114(30.2) | 90(23.9) | <0.001 |
| Age≤45 Yr | 105(27.9) | 72(19.1) | 58(15.4) | <0.001 |
| Age, 46-60 Yr | 131(34.7) | 145(38.5) | 122(32.4) | 0.041 |
| Age>60 Yr | 141(37.4) | 160(42.4) | 196(52.1) | <0.001 |
| **Medical comorbidities** |  |  |  |  |
| Hypertension | 161(42.7) | 170(45.1) | 175(46.5) | 0.565 |
| Cardiovascular disease | 63(16.7) | 58(15.4) | 78(20.7) | 0.147 |
| Diabetes | 131(34.7) | 163(43.2) | 164(43.6) | 0.020 |
| Chronic obstructive pulmonary disease | 08(2.1) | 08(2.1) | 11(2.9) | 0.474 |
| **Clinical features & investigations** |  |  |  |  |
| Fever | 295(78.2) | 322(85.4) | 327(87.0) | 0.001 |
| Shortness of breath | 126(33.4) | 168(44.6) | 225(59.8) | <0.001 |
| Pulse (per minute) | 86.0(78.0-97.5) | 88.0(79.0-99.5) | 93.0(82.0-105.0) | <0.001 |
| Oxygen (SpO<sub>2</sub> %) | 97.0(95.0-98.0) | 95.0(92.0-97.0) | 91.0(82.2-95.0) | <0.001 |
| Systolic BP mmHg | 130(121.5-142.0) | 130.0(120.0-142.0) | 132.0(120.0-148.0) | 0.314 |
| Hemoglobin (g/dl) | 12.8(11.7-14.1) | 12.8(11.5-14.0) | 12.6(11.1-13.8) | 0.019 |
| White cell count (10<sup>3</sup>/μl) | 5.8(4.5-7.6) | 6.8(5.1-9.5) | 8.2(5.9-11.9) | <0.001 |
| Neutrophils (%) | 71.3(60.6-79.8) | 78.9(70.7-86.7) | 86.9(79.6-91.9) | <0.001 |
| Lymphocytes (%) | 23.4(15.5-32.5) | 15.7(9.5-23.7) | 9.6(5.9-16.2) | <0.001 |
| Platelet count (10<sup>6</sup>/μl) | 2.2(1.7-2.7) | 2.1(1.7-2.7) | 2.2(1.6-2.9) | 0.807 |
| d-Dimer (ng/dl) | 348.5(167.0-835.2) | 421.0(190.5-744.0) | 602.3(268.5-1454.7) | <0.001 |
| Interleukin-6 (ng/dl) | 8.3(2.9-22.7) | 21.1(6.8-53.2) | 45.8(16.5-117.3) | <0.001 |
| Ferritin (mg/dl) | 190.1(101.0-365.3) | 356.9(171.4-655.1) | 530.0(296.1-1008.5) | <0.001 |
| Lactic dehydrogenase (mg/dl) | 213.0(181.0-265.0) | 283.0(236.0-376.0) | 368.0(284.0-516.3) | <0.001 |
| Creatinine (mg/dl) | 0.87(0.74-1.08) | 0.92(0.77-1.12) | 1.03(0.84-1.42) | <0.001 |
| Sodium (mEq/L) | 139.0(137.0-142.0) | 138.0(135.0-141.0) | 137.0(134.0-141.0) | <0.001 |
| Potassium (mEq/L) | 4.3(4.0-4.7) | 4.4(4.1-4.8) | 4.6(4.1-5.0) | <0.001 |
| Serum aspartate transaminase (units) | 25.0(17.7-39.7) | 29.1(19.3-47.5) | 31.0(19.4-48.8) | 0.001 |
| Serum glutamate transaminase (units) | 25.2(19.0-36.3) | 32.5(22.7-50.5) | 35.7(25.3-53.9) | <0.001 |
| HRCT scan thorax score (out of 25) | 9(4.0-12.0) | 13.0(9.5-17.0) | 16.0(11.0-19.0) | <0.001 |
| **Medicines** |  |  |  |  |
| Steroids | 343(91.0) | 363(96.3) | 366(97.3) | <0.001 |
| Remdesivir | 301(79.8) | 344(91.2) | 349(92.8) | <0.001 |
| Anticoagulants | 346(91.8) | 359(95.2) | 354(94.1) | 0.180 |
| Tocilizumab/ Bevacizumab | 4(1.1) | 11(2.9) | 57(13.2) | <0.001 |
| **Outcomes** |  |  |  |  |
| Oxygen requirement at admission | 50(13.3) | 135(25.8) | 252(67.0) | <0.001 |
| Oxygen duration (days) | 0(0.0-0.0) | 0.0(0.0-5.0) | 6(2-11) | <0.001 |
| Proning | 137(36.7) | 226(59.9) | 301(80.1) | <0.001 |
| High flow nasal cannula | 9(2.4) | 29(7.7) | 97(25.8) | <0.001 |
| BiPaP support | 8(2.1) | 26(6.9) | 95(25.3) | <0.001 |
| Invasive ventilation | 8(2.1) | 21(5.6) | 101(26.9) | <0.001 |
| Total length of stay (day) | 7.0(6.0-9.0) | 8.0(6.0-8.0) | 9.0(7.0-12.7) | <0.001 |
| ICU (days) | 7.0(3.0-9.2) | 7.0(5.0-10.0) | 9.0(5.0-14.0) | 0.003 |
| Transfer to hospice care | 24(6.4) | 39(10.3) | 61(16.2) | <0.001 |
| Deaths | 12(3.2) | 27(7.2) | 117(31.1) | <0.001 |

**Table 2:** D-dimer tertiles, clinical features and outcomes.

|  | Tertile 1<br>(N=287, ≤266) | Tertile 2<br>(N=287, 266-720) | Tertile 3<br>(N=287, ≥721) | Statistics |
| --- | --- | --- | --- | --- |
| Male | 196(68.3) | 182(63.4) | 209(72.8) | 0.053 |
| Female | 91(31.7) | 105(36.6) | 78(27.2) | 0.072 |
| Age≤45 Yr | 67(23.3) | 49(17.1) | 41(14.3) | <0.001 |
| Age, 46-60 Yr | 125(43.6) | 111(38.7) | 76(26.5) | <0.001 |
| Age>60 Yr | 95(33.1) | 127(44.3) | 170(59.2) | <0.001 |
| **Medical comorbidities** |  |  |  |  |
| Hypertension | 112(39.0) | 142(49.5) | 140(48.8) | 0.019 |
| Cardiovascular disease | 42(14.6) | 43(15.0) | 69(24.0) | 0.003 |
| Diabetes | 102(35.5) | 121(42.2) | 125(43.6) | 0.051 |
| Chronic obstructive pulmonary disease | 02(0.7) | 03(1.0) | 11(3.8) | 0.010 |
| **Clinical features & investigations** |  |  |  |  |
| Fever | 252(87.8) | 252(87.8) | 235(81.9) | 0.042 |
| Shortness of breath | 100(34.8) | 126(43.9) | 177(61.7) | <0.001 |
| Pulse (per minute) | 88.0(80.0-100.0) | 89.0(80.0-100.0) | 90.0(80.0-104.0) | 0.462 |
| Oxygen (SpO<sub>2</sub> %) | 96.0(93.0-97.0) | 95.0(91.0-97.0) | 92.0(83.0-96.0) | <0.001 |
| Systolic BP mmHg | 131.0(120.0-144.0) | 134.0(123.5-150.0) | 130.0(120.0-143.0) | 0.099 |
| Hemoglobin (g/dl) | 13.3(12.2-14.4) | 12.7(11.6-13.9) | 12.1(10.7-13.5) | <0.001 |
| White cell count (10<sup>3</sup>/μl) | 6.5(4.8-8.6) | 6.5(5.0-9.2) | 7.8(5.7-12.0) | <0.001 |
| Neutrophils (%) | 77.4(64.3-85.6) | 79.0(69.2-87.6) | 85.6(76.5-91.5) | <0.001 |
| Lymphocytes (%) | 17.6(11.3-30.0) | 15.7(9.2-24.8) | 11.0(5.9-19.6) | <0.001 |
| Platelet count (10<sup>6</sup>/μl) | 2.3(1.7-2.8) | 2.1(1.7-2.7) | 2.2(1.8-2.9) | 0.349 |
| hs-CRP (mg/dl) | 5.9(2.1-16.3) | 9.0(3.8-20.5) | 13.3(4.1-34.1) | <0.001 |
| Interleukin-6 (ng/dl) | 13.4(3.6-36.3) | 20.3(7.8-53.2) | 36.1(14.3-113.2) | <0.001 |
| Ferritin (mg/dl) | 252.9(118.5-560.8) | 339.6(178.3-649.4) | 481.8(212.3-993.0) | <0.001 |
| Lactic dehydrogenase (mg/dl) | 258.0(208.0-314.0) | 290.0(228.7-390.0) | 369.0(252.2-545.2) | <0.001 |
| Creatinine (mg/dl) | 0.89(0.77-1.06) | 0.93(0.80-1.20) | 1.0(0.79-1.40) | <0.001 |
| Sodium (mEq/L) | 139.0(135.5-141.0) | 138.0(135.0-141.0) | 138.0(134.0-142.0) | 0.805 |
| Potassium (mEq/L) | 4.5(4.2-4.8) | 4.4(4.1-4.8) | 4.5(4.1-4.9) | 0.861 |
| Serum aspartate transaminase (units) | 27.5(18.8-42.8) | 28.3(18.4-46.4) | 28.2(19.4-46.6) | 0.622 |
| Serum glutamate transaminase (units) | 28.1(20.2-39.9) | 32.0(22.0-47.4) | 34.2(25.2-57.8) | <0.001 |
| HRCT scan thorax score (out of 25) | 11.0(7.2-15.0) | 12.0(8.0-17.0) | 15.5(10.0-20.0) | <0.001 |
| **Medicines** |  |  |  |  |
| Steroids | 276(96.2) | 280(97.6) | 276(96.2) | 1.00 |
| Remdesivir | 266(92.7) | 274(95.5) | 260(90.6) | 0.329 |
| Anticoagulants | 277(96.5) | 279(97.2) | 270(94.1) | 0.139 |
| Tocilizumab/ Bevacizumab | 06(2.1) | 13(4.5) | 41(14.3) | <0.001 |
| **Outcomes** |  |  |  |  |
| O<sub>2</sub> required | 69(24.0) | 110(38.3) | 182(63.4) | <0.001 |
| Duration (days) | 0.0(0.0-4.0) | 2(0.0-6.0) | 5.0(0.0-9.0) | <0.001 |
| Proning | 204(71.1) | 206(71.8) | 196(68.3) | 0.626 |
| Nasal cannula | 66(23.0) | 94(32.8) | 120(41.8) | <0.001 |
| High flow nasal | 19(6.6) | 36(12.5) | 54(18.8) | <0.001 |
| BiPaP support | 16(5.6) | 31(10.8) | 60(20.9) | <0.001 |
| Invasive ventilation | 09(3.1) | 23(8.0) | 78(27.2) | <0.001 |
| Total length of stay (day) | 7.0(6.0-9.0) | 8.0(6.0-10.0) | 9.0(7.0-12.0) | <0.001 |
| ICU (days) | 9.0(5.0-12.0) | 8.0(5.0-14.0) | 8.0(5.7-13.0) | 0.002 |
| Transfer to hospice care | 11(3.8) | 25(8.7) | 43(15.0) | <0.001 |
| Deaths | 09(3.1) | 28(9.8) | 91(31.7) | <0.001 |

**Table 3:** Interleukin-6 (IL-6) tertiles, clinical features and outcomes.

|  | Tertile 1<br>(N=346, ≤10.3) | Tertile 2<br>(N=346, 10.4-39.7) | Tertile 3<br>(N=346, ≥39.8) | Statistics |
| --- | --- | --- | --- | --- |
| Male | 224(64.7) | 250(72.3) | 259(74.9) | 0.004 |
| Female | 122(35.3) | 96(27.7) | 87(25.1) | <0.001 |
| Age≤45 Yr | 92(26.6) | 77(22.3) | 41(11.8) | <0.001 |
| Age, 46-60 Yr | 132(38.2) | 123(35.5) | 123(35.5) | <0.001 |
| Age>60 Yr | 122(35.3) | 146(42.2) | 182(52.6) | <0.001 |
| **Medical comorbidities** |  |  |  |  |
| Hypertension | 138(39.9) | 152(43.9) | 176(50.9) | 0.004 |
| Cardiovascular disease | 48(13.9) | 58(16.8) | 73(21.1) | 0.012 |
| Diabetes | 122(35.3) | 134(38.7) | 164(47.4) | 0.001 |
| Chronic obstructive pulmonary disease | 07(2.0) | 06(1.7) | 11(3.2) | 0.408 |
| **Clinical features & investigations** |  |  |  |  |
| Fever | 283(81.8) | 306(88.4) | 297(85.8) | 0.045 |
| Shortness of breath | 125(36.1) | 149(43.1) | 194(56.1) | <0.001 |
| Pulse (per minute) | 86.0(78.0-97.0) | 89.0(80.0-100.0) | 92.0(82.0-106.0) | <0.001 |
| Oxygen (SpO<sub>2</sub> %) | 96.0(94.0-98.0) | 95.0(92.0-97.0) | 92.5(85.0-96.0) | <0.001 |
| Systolic BP mmHg | 131.0(122.7-145.0) | 131.0(122.0-142.2) | 130.0(120.0-144.0) | 0.418 |
| Hemoglobin (g/dl) | 13.0(11.8-14.1) | 12.9(11.8-14.1) | 12.4(11.1-13.8) | <0.001 |
| White cell count (10<sup>3</sup>/μl) | 6.0(4.6-8.6) | 6.5(4.7-8.9) | 8.0(6.0-11.6) | <0.001 |
| Neutrophils (%) | 75.5(64.8-84.7) | 78.4(67.8-88.5) | 83.7(75.2-90.5) | <0.001 |
| Lymphocytes (%) | 19.3(11.6-28.7) | 17.1(8.5-27.9) | 12.1(7.0-19.0) | <0.001 |
| Platelet count (10<sup>6</sup>/μl) | 2.4(1.9-2.9) | 2.1(1.6-2.7) | 2.1(1.6-2.7) | <0.001 |
| hs-CRP (mg/dl) | 3.0(0.89-9.2) | 7.0(2.7-17.7) | 15.3(6.8-35.5) | <0.001 |
| d-Dimer (ng/dl) | 306.6(167.0-614.9) | 450.0(184.1-869.5) | 622.5(292.7-1368.7) | <0.001 |
| Ferritin (mg/dl) | 230.9(102.1-504.4) | 348.6(173.8-614.1) | 477.1(244.2-884.2) | <0.001 |
| Lactic dehydrogenase (mg/dl) | 241.5(192.0-320.0) | 276.5(224.0-375.7) | 345.5(264.2-499.7) | <0.001 |
| Creatinine (mg/dl) | 0.87-0.75-1.06() | 0.94(0.78-1.19) | 1.02(0.83-1.42) | <0.001 |
| Sodium (mEq/L) | 140(136.5-142.0) | 138.0(135.0-141.0) | 137.0(134.0-140.0) | <0.001 |
| Potassium (mEq/L) | 4.5(4.1-4.8) | 4.4(4.1-4.8) | 4.4(4.0-4.9) | 0.663 |
| Serum aspartate transaminase (units) | 28.1(18.8-44.0) | 26.6(17.5-43.8) | 29.7(19.5-47.9) | 0.440 |
| Serum glutamate transaminase (units) | 26.7(19.6-39.6) | 30.1(21.5-47.5) | 37.8(25.0-57.4) | <0.001 |
| HRCT scan thorax score (out of 25) | 10.0(4.2-14.0) | 12.0(7.0-16.0) | 15.0(12.0-19.0) | <0.001 |
| **Medicines** |  |  |  |  |
| Steroids | 332(96.0) | 338(97.7) | 336(97.7) | 0.379 |
| Remdesivir | 307(88.7) | 322(93.1) | 321(92.8) | 0.056 |
| Anticoagulants | 329(95.1) | 333(96.2) | 325(93.9) | 0.372 |
| Tocilizumab/ Bevacizumab | 04(1.2) | 12(3.5) | 54(15.6) | <0.001 |
| **Outcomes** |  |  |  |  |
| O<sub>2</sub> required | 68(19.7) | 122(35.3) | 219(63.3) | <0.001 |
| Duration (days) | 0.0(0.0-0.0) | 0.0(0.0-6.0) | 5.0(0.0-9.0) | <0.001 |
| Proning | 179(51.7) | 220(63.6) | 242(69.9) | <0.001 |
| Nasal cannula | 60(17.3) | 109(31.5) | 150(43.4) | <0.001 |
| High flow nasal | 16(4.6) | 40(11.6) | 73(21.1) | <0.001 |
| BiPaP support | 15(4.3) | 32(9.2) | 76(22.0) | <0.001 |
| Invasive ventilation | 13(3.8) | 23(6.6) | 86(24.9) | <0.001 |
| Length of stay (day) | 7.0(6.0-9.0) | 8.0(6.0-10.0) | 9.0(7.0-12.0) | <0.001 |
| ICU (days) | 7.0(4.0-10.2) | 8.0(6.0-12.5) | 8.0(5.0-13.7) | 0.639 |
| Transfer to hospice care | 25(7.2) | 31(9.0) | 50(14.3) | <0.001 |
| Deaths | 12(3.5) | 30(8.7) | 100(28.9) | <0.001 |

**Table 4:** Lactic dehydrogenase (LDH) tertiles, clinical features and outcomes.

|  | Tertile 1<br>(N=241, ≤242) | Tertile 2<br>(N=240, 243-347) | Tertile 3<br>(N=240, ≥348) | Statistics |
| --- | --- | --- | --- | --- |
| Male | 149(61.8) | 181(75.4) | 170(70.8) | 0.032 |
| Female | 92(38.2) | 59(24.6) | 70(29.2) | 0.043 |
| Age≤45 Yr | 64(26.6) | 42(17.5) | 35(14.6) | <0.001 |
| Age, 46-60 Yr | 87(36.1) | 98(40.8) | 95(39.6) | 0.236 |
| Age>60 Yr | 90(37.3) | 100(41.7) | 110(45.8) | <0.001 |
| **Medical comorbidities** |  |  |  |  |
| Hypertension | 98(40.7) | 119(49.6) | 110(45.8) | 0.254 |
| Cardiovascular disease | 41(17.0) | 41(17.1) | 36(15.0) | 0.551 |
| Diabetes | 94(39.0) | 102(42.5) | 92(38.3) | 0.882 |
| Chronic obstructive pulmonary disease | 08(3.3) | 02(0.8) | 06(2.5) | 0.167 |
| **Clinical features & investigations** |  |  |  |  |
| Fever | 195(80.9) | 210(87.5) | 205(85.4) | 0.177 |
| Shortness of breath | 69(28.6) | 108(45.0) | 167(69.6) | <0.001 |
| Pulse (per minute) | 86.0(78.0-98.5) | 90.0(81.0-101.7) | 90.0(80.0-104.7) | 0.135 |
| Oxygen (SpO<sub>2</sub> %) | 97.0(95.0-98.0) | 96.0(92.0-97.0) | 90.0(80.0-94.0) | <0.001 |
| Systolic BP mmHg | 130.0(120.0-141.5) | 133.0(122.2-146.0) | 133.0(122.0-146.0) | 0.082 |
| Hemoglobin (g/dl) | 12.8(11.6-14.0) | 12.8(11.6-14.2) | 12.8(11.5-13.9) | 0.762 |
| White cell count (10<sup>3</sup>/μl) | 5.8(4.5-7.3) | 6.7(4.7-9.7) | 8.4(5.9-12.1) | <0.001 |
| Neutrophils (%) | 71.9(59.8-81.3) | 79.2(70.6-88.2) | 87.0(80.0-91.5) | <0.001 |
| Lymphocytes (%) | 22.1(14.2-32.2) | 14.9(8.5-23.4) | 9.4(6.0-16.5) | <0.001 |
| Platelet count (10<sup>6</sup>/μl) | 2.2(1.7-2.8) | 2.2(1.6-2.8) | 2.1(1.6-2.9) | 0.729 |
| hs-CRP (mg/dl) | 2.3(0.75-6.7) | 8.8(4.2-20.0) | 18.2(8.4-39.6) | <0.001 |
| d-Dimer (ng/dl) | 292.0(167.0-633.1) | 346.5(182.7-668.1) | 758.4(382.2-1715.1) | <0.001 |
| Interleukin-6 (ng/dl) | 10.8(3.8-28.1) | 25.3(9.6-66.2) | 42.8(13.1-112.3) | <0.001 |
| Ferritin (mg/dl) | 180.9(84.3-320.7) | 357.2(169.3-618.9) | 592.0(374.0-1173.0) | <0.001 |
| Creatinine (mg/dl) | 0.88(0.75-1.08) | 0.94(0.78-1.14) | 1.01(0.84-1.37) | 0.001 |
| Sodium (mEq/L) | 139.0(137.0-141.0) | 137.0(134.0-140.0) | 138.0(134.0-141.0) | <0.001 |
| Potassium (mEq/L) | 4.3(4.0-4.7) | 4.5(4.1-4.8) | 4.6(4.1-5.1) | 0.002 |
| Serum aspartate transaminase (units) | 21.6(15.4-30.7) | 32.9(21.0-48.6) | 37.0(23.1-63.1) | <0.001 |
| Serum glutamate transaminase (units) | 22.5(17.4-29.6) | 32.6(23.6-46.0) | 45.6(31.5-68.7) | <0.001 |
| HRCT scan thorax score (out of 25) | 7.0(2.0-11.0) | 13.0(10.0-16.0) | 17.0(13.2-21.0) | <0.001 |
| **Medicines** |  |  |  |  |
| Steroids | 214(88.8) | 236(98.3) | 236(98.3) | <0.001 |
| Remdesivir | 199(82.6) | 226(94.2) | 221(92.1) | 0.001 |
| Anticoagulants | 216(89.6) | 229(95.4) | 233(97.1) | 0.001 |
| Tocilizumab/ Bevacizumab | 03(1.2) | 13(5.4) | 42(17.5) | <0.001 |
| **Outcomes** |  |  |  |  |
| O<sub>2</sub> required | 42(17.4) | 85(35.4) | 178(74.2) | <0.001 |
| Duration (days) | 0.0(0.0-0.0) | 0.0(0.0-6.0) | 6.0(2.0-10.5) | <0.001 |
| Proning | 116(48.1) | 151(62.9) | 190(79.2) | <0.001 |
| Nasal cannula | 39(16.2) | 78(32.5) | 119(49.6) | <0.001 |
| High flow nasal | 10(4.1) | 19(7.9) | 66(27.5) | <0.001 |
| BiPaP support | 07(2.9) | 19(7.9) | 68(28.3) | <0.001 |
| Invasive ventilation | 08(3.3) | 15(6.3) | 70(29.2) | <0.001 |
| Length of stay (day) | 7.0(6.0-8.0) | 8.0(6.0-10.7) | 9.0(7.0-13.0) | <0.001 |
| ICU (days) | 8.0(4.0-10.0) | 9.0(6.0-15.0) | 8.0(5.0-14.0) | 0.265 |
| Transfer to hospice care | 27(11.2) | 17(7.1) | 46(19.2) | 0.237 |
| Deaths | 11(4.6) | 20(8.3) | 80(33.3) | <0.001 |

**Table 5:**
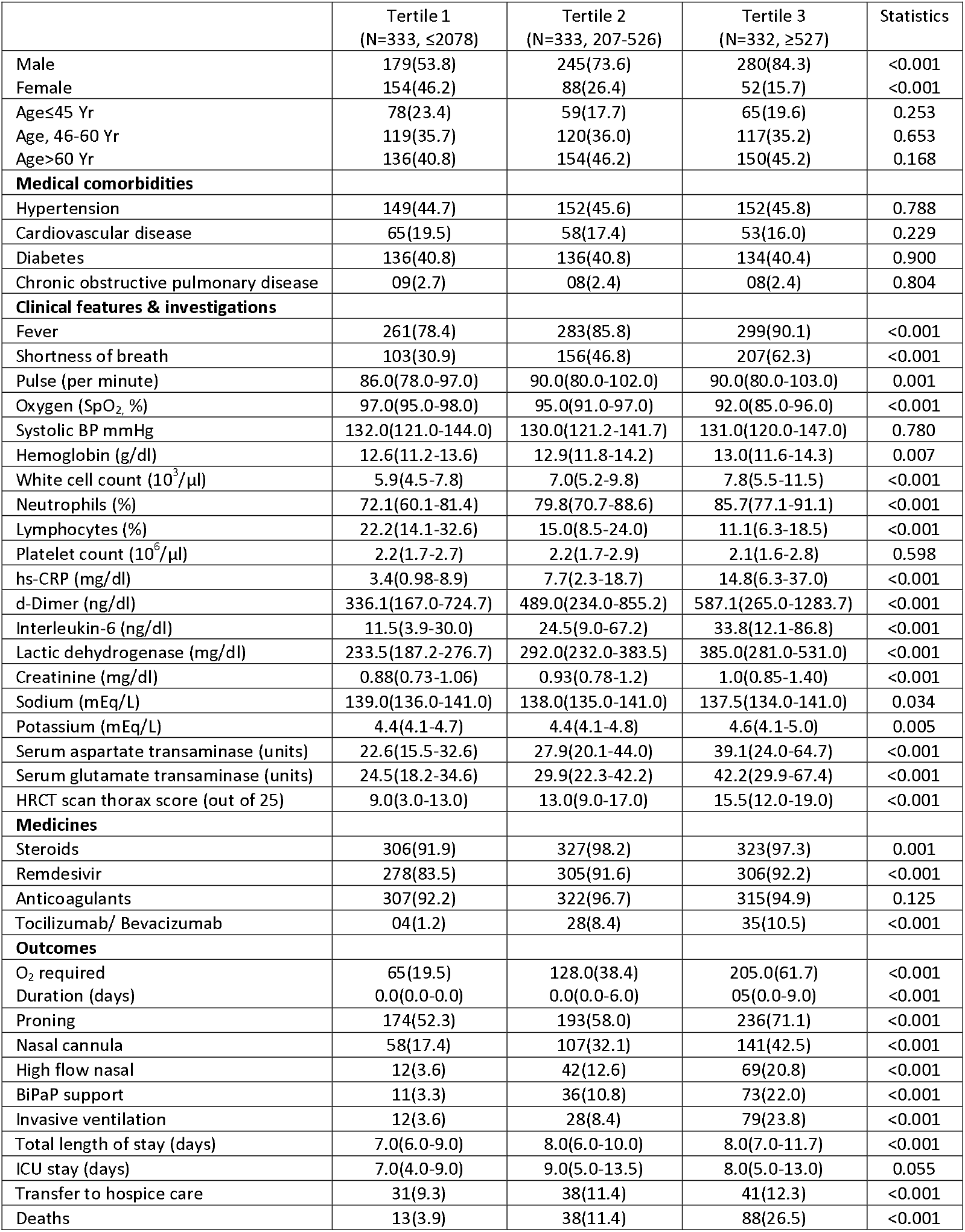
Ferritin tertiles, clinical features and outcomes.

|  | Tertile 1<br>(N=333, ≤2078) | Tertile 2<br>(N=333, 207-526) | Tertile 3<br>(N=332, ≥527) | Statistics |
| --- | --- | --- | --- | --- |
| Male | 179(53.8) | 245(73.6) | 280(84.3) | <0.001 |
| Female | 154(46.2) | 88(26.4) | 52(15.7) | <0.001 |
| Age≤45 Yr | 78(23.4) | 59(17.7) | 65(19.6) | 0.253 |
| Age, 46-60 Yr | 119(35.7) | 120(36.0) | 117(35.2) | 0.653 |
| Age>60 Yr | 136(40.8) | 154(46.2) | 150(45.2) | 0.168 |
| **Medical comorbidities** |  |  |  |  |
| Hypertension | 149(44.7) | 152(45.6) | 152(45.8) | 0.788 |
| Cardiovascular disease | 65(19.5) | 58(17.4) | 53(16.0) | 0.229 |
| Diabetes | 136(40.8) | 136(40.8) | 134(40.4) | 0.900 |
| Chronic obstructive pulmonary disease | 09(2.7) | 08(2.4) | 08(2.4) | 0.804 |
| **Clinical features & investigations** |  |  |  |  |
| Fever | 261(78.4) | 283(85.8) | 299(90.1) | <0.001 |
| Shortness of breath | 103(30.9) | 156(46.8) | 207(62.3) | <0.001 |
| Pulse (per minute) | 86.0(78.0-97.0) | 90.0(80.0-102.0) | 90.0(80.0-103.0) | 0.001 |
| Oxygen (SpO<sub>2</sub> %) | 97.0(95.0-98.0) | 95.0(91.0-97.0) | 92.0(85.0-96.0) | <0.001 |
| Systolic BP mmHg | 132.0(121.0-144.0) | 130.0(121.2-141.7) | 131.0(120.0-147.0) | 0.780 |
| Hemoglobin (g/dl) | 12.6(11.2-13.6) | 12.9(11.8-14.2) | 13.0(11.6-14.3) | 0.007 |
| White cell count (10<sup>3</sup>/μl) | 5.9(4.5-7.8) | 7.0(5.2-9.8) | 7.8(5.5-11.5) | <0.001 |
| Neutrophils (%) | 72.1(60.1-81.4) | 79.8(70.7-88.6) | 85.7(77.1-91.1) | <0.001 |
| Lymphocytes (%) | 22.2(14.1-32.6) | 15.0(8.5-24.0) | 11.1(6.3-18.5) | <0.001 |
| Platelet count (10<sup>6</sup>/μl) | 2.2(1.7-2.7) | 2.2(1.7-2.9) | 2.1(1.6-2.8) | 0.598 |
| hs-CRP (mg/dl) | 3.4(0.98-8.9) | 7.7(2.3-18.7) | 14.8(6.3-37.0) | <0.001 |
| d-Dimer (ng/dl) | 336.1(167.0-724.7) | 489.0(234.0-855.2) | 587.1(265.0-1283.7) | <0.001 |
| Interleukin-6 (ng/dl) | 11.5(3.9-30.0) | 24.5(9.0-67.2) | 33.8(12.1-86.8) | <0.001 |
| Lactic dehydrogenase (mg/dl) | 233.5(187.2-276.7) | 292.0(232.0-383.5) | 385.0(281.0-531.0) | <0.001 |
| Creatinine (mg/dl) | 0.88(0.73-1.06) | 0.93(0.78-1.2) | 1.0(0.85-1.40) | <0.001 |
| Sodium (mEq/L) | 139.0(136.0-141.0) | 138.0(135.0-141.0) | 137.5(134.0-141.0) | 0.034 |
| Potassium (mEq/L) | 4.4(4.1-4.7) | 4.4(4.1-4.8) | 4.6(4.1-5.0) | 0.005 |
| Serum aspartate transaminase (units) | 22.6(15.5-32.6) | 27.9(20.1-44.0) | 39.1(24.0-64.7) | <0.001 |
| Serum glutamate transaminase (units) | 24.5(18.2-34.6) | 29.9(22.3-42.2) | 42.2(29.9-67.4) | <0.001 |
| HRCT scan thorax score (out of 25) | 9.0(3.0-13.0) | 13.0(9.0-17.0) | 15.5(12.0-19.0) | <0.001 |
| **Medicines** |  |  |  |  |
| Steroids | 306(91.9) | 327(98.2) | 323(97.3) | 0.001 |
| Remdesivir | 278(83.5) | 305(91.6) | 306(92.2) | <0.001 |
| Anticoagulants | 307(92.2) | 322(96.7) | 315(94.9) | 0.125 |
| Tocilizumab/ Bevacizumab | 04(1.2) | 28(8.4) | 35(10.5) | <0.001 |
| **Outcomes** |  |  |  |  |
| O<sub>2</sub> required | 65(19.5) | 128.0(38.4) | 205.0(61.7) | <0.001 |
| Duration (days) | 0.0(0.0-0.0) | 0.0(0.0-6.0) | 05(0.0-9.0) | <0.001 |
| Proning | 174(52.3) | 193(58.0) | 236(71.1) | <0.001 |
| Nasal cannula | 58(17.4) | 107(32.1) | 141(42.5) | <0.001 |
| High flow nasal | 12(3.6) | 42(12.6) | 69(20.8) | <0.001 |
| BiPaP support | 11(3.3) | 36(10.8) | 73(22.0) | <0.001 |
| Invasive ventilation | 12(3.6) | 28(8.4) | 79(23.8) | <0.001 |
| Total length of stay (days) | 7.0(6.0-9.0) | 8.0(6.0-10.0) | 8.0(7.0-11.7) | <0.001 |
| ICU stay (days) | 7.0(4.0-9.0) | 9.0(5.0-13.5) | 8.0(5.0-13.0) | 0.055 |
| Transfer to hospice care | 31(9.3) | 38(11.4) | 41(12.3) | <0.001 |
| Deaths | 13(3.9) | 38(11.4) | 88(26.5) | <0.001 |

**Table 6:** Neutrophil:lymphocyte ratio tertiles, clinical features and outcomes.

|  | Tertile 1<br>(N=405, ≤3.34) | Tertile 2<br>(N=405, 3.35-7.75) | Tertile 3<br>(N=404, ≥7.76) | Statistics |
| --- | --- | --- | --- | --- |
| Male | 255(63.0) | 276(68.1) | 319(79.0) | <0.001 |
| Female | 150(37.0) | 129(31.9) | 85(21.0) | <0.001 |
| Age≤45 Yr | 112(27.7) | 78(19.3) | 62(15.3) | <0.001 |
| Age, 46-60 Yr | 134(33.1) | 159(39.3) | 130(32.2) | 0.346 |
| Age>60 Yr | 159(39.3) | 168(41.5) | 212(52.5) | <0.001 |
| **Medical comorbidities** |  |  |  |  |
| Hypertension | 158(39.0) | 181(44.7) | 197(48.8) | 0.005 |
| Cardiovascular disease | 71(17.5) | 71(17.5) | 86(21.3) | 0.172 |
| Diabetes | 142(35.1) | 169(41.7) | 171(42.3) | 0.035 |
| Chronic obstructive pulmonary disease | 09(2.2) | 14(3.5) | 09(2.2) | 1.00 |
| **Clinical features & investigations** |  |  |  |  |
| Fever | 320(79.0) | 323(79.8) | 324(80.2) | 0.675 |
| Shortness of breath | 118(29.1) | 176(43.5) | 252(62.5) | <0.001 |
| Pulse (per minute) | 85.0(78.0-95.0) | 90.0(80.0-100.0) | 91.0(80.0-104.7) | <0.001 |
| Oxygen (SpO<sub>2</sub> %) | 97.0(95.0-98.0) | 95.0(92.0-97.0) | 91.0(82.0-95.0) | <0.001 |
| Systolic BP mmHg | 130.0(120.0-140.0) | 130.0(120.0-142.0) | 133.0(120.0-148.0) | 0.145 |
| Hemoglobin (g/dl) | 12.8(11.8-14.0) | 12.7(11.3-14.0) | 12.6(11.1-13.9) | 0.086 |
| White cell count (10<sup>3</sup>/μl) | 5.3(4.2-6.8) | 6.7(5.3-8.8) | 10.0(7.4-13.6) | <0.001 |
| Neutrophils (%) | 63.4(56.1-68.5) | 79.5(76.4-82.6) | 90.8(88.1-93.4) | <0.001 |
| Lymphocytes (%) | 30.4(25.5-35.9) | 15.9(13.4-18.9) | 6.8(4.4-8.8) | <0.001 |
| Platelet count (10<sup>6</sup>/μl) | 2.1(1.7-2.7) | 2.2(1.8-2.8) | 2.2(1.6-2.9) | 0.487 |
| hs-CRP (mg/dl) | 2.6(0.82-7.3) | 6.8(2.6-16.0) | 16.6(6.7-36.6) | <0.001 |
| d-Dimer (ng/dl) | 329.8(167.0-618.7) | 428.5(187.3-806.8) | 662.0(294.0-1517.0) | <0.001 |
| Interleukin-6 (ng/dl) | 13.8(4.0-36.2) | 19.8(5.9-54.8) | 30.3(11.9-102.8) | <0.001 |
| Ferritin (mg/dl) | 196.1(113.1-416.2) | 333.9(169.8-709.9) | 523.3(314.4-996.0) | <0.001 |
| Lactic dehydrogenase (mg/dl) | 236(188.5-285.0) | 277.0(219.0-376.0) | 368.0(281.0-518.0) | <0.001 |
| Creatinine (mg/dl) | 0.87(0.74-1.04) | 0.95(0.76-1.20) | 1.0(0.86-1.42) | <0.001 |
| Sodium (mEq/L) | 140.0(137.0-142.0) | 138.0(134.0-141.0) | 137.5(134.0-141.0) | <0.001 |
| Potassium (mEq/L) | 4.4(4.1-4.7) | 4.4(4.1-4.8) | 4.5(4.1-5.0) | 0.001 |
| Serum aspartate transaminase (units) | 25.7(17.4-40.1) | 26.8(17.4-43.5) | 32.7(21.0-53.8) | 0.007 |
| Serum glutamate transaminase (units) | 27.1(19.7-41.0) | 30.5(21.3-45.1) | 35.4(22.7-55.4) | <0.001 |
| HRCT scan thorax score (out of 25) | 10.0(4.0-12.0) | 12.0(8.0-17.0) | 16.0(12.0-20.0) | <0.001 |
| **Medicines** |  |  |  |  |
| Steroids | 352(86.9) | 362(89.4) | 375(92.8) | 0.006 |
| Remdesivir | 315(77.8) | 334(82.5) | 351(86.9) | 0.001 |
| Anticoagulants | 354(87.4) | 367(90.6) | 363(89.9) | 0.261 |
| Tocilizumab/ Bevacizumab | 02(0.5) | 18(4.4) | 53(13.1) | <0.001 |
| **Outcomes** |  |  |  |  |
| O<sub>2</sub> required | 57(14.1) | 136(33.6) | 275(68.1) | <0.001 |
| Duration (days) | 0(0.0) | 0.0(0.0-5.0) | 5.0(1.0-9.0) | <0.001 |
| Proning | 169(41.7) | 218(53.8) | 283(70.0) | <0.001 |
| Nasal cannula | 50(12.3) | 116(28.6) | 184(45.5) | <0.001 |
| High flow nasal | 05(1.2) | 37(9.1) | 96(23.8) | <0.001 |
| BiPaP support | 09(2.2) | 29(7.2) | 95(23.5) | <0.001 |
| Invasive ventilation | 08(2.0) | 27(6.7) | 107(26.5) | <0.001 |
| Length of stay (day) | 7.0(5.0-8.0) | 8.0(6.0-9.0) | 8.0(6.0-12.0) | <0.001 |
| ICU (days) | 5.0(2.0-8.0) | 7.0(3.7-10.0) | 8.0(4.0-13.0) | 0.002 |
| Transfer to hospice care | 50(12.3) | 57(14.1) | 78(19.3) | <0.001 |
| Deaths | 10(2.5) | 33(8.1) | 128(31.7) | <0.001 |

**Table 7:** Correlation matrix (parametric Pearson’s r and non-parametric Spearman’s rho) among the biomarkers evaluated.

| Variable | hsCRP | d-Dimer | Interleukin-6 | Lactic dehydrogenase | Ferritin | Neutrophil-lymphocyte ratio |
| --- | --- | --- | --- | --- | --- | --- |
| **Pearson correlation (r)** |  |  |  |  |  |  |
| hsCRP | --- | 0.180** | 0.142** | 0.287** | 0.269** | 0.305** |
| d-Dimer | 0.180** | --- | 0.262** | 0.346** | 0.224** | 0.225** |
| Interleukin-6 | 0.142** | 0.262** | --- | 0.317** | 0.166** | 0.202** |
| Lactic dehydrogenase | 0.287** | 0.346** | 0.377** | --- | 0.416** | 0.308** |
| Ferritin | 0.269** | 0.224** | 0.166** | 0.416** | --- | 0.256** |
| Neutrophil-lymphocyte | 0.305** | 0.225** | 0.202** | 0.308** | 0.256** | --- |
| **Spearman correlation (rho)** |  |  |  |  |  |  |
| hsCRP | --- | 0.231** | 0.424** | 0.543** | 0.420** | 0.477** |
| d-Dimer | 0.231** | --- | 0.288** | 0.357** | 0.227** | 0.282** |
| Interleukin-6 | 0.424** | 0.288** | --- | 0.360** | 0.280** | 0.266** |
| Lactic dehydrogenase | 0.543** | 0.357** | 0.360** | --- | 0.521** | 0.477** |
| Ferritin | 0.420** | 0.227** | 0.280** | 0.521** | --- | 0.383** |
| Neutrophil-lymphocyte | 0.477** | 0.282** | 0.266** | 0.477** | 0.383** | --- |

Incidence of deaths in various biomarker groups is shown in Figure 1. For all the biomarkers, incidence of death is very high in the highest tertile. Univariate, age and sex adjusted and multivariate (age, sex, risk factors, comorbidities) adjusted odds ratio (95% CI) for deaths in second and third vs first tertiles are in Table 8. Figure 2 shows the incidence of multivariate adjusted OR (95% CI) for in-hospital deaths in various biomarker groups. As compared to low mortality in the lowest tertile (reference group), deaths were significantly greater in second and third tertiles for all the biomarkers. Multivariate ORs (95% CI) in second and third tertiles, compared to the lowest, respectively, were for, hsCRP 2.29 (1.14-4.60) and 13.39 (7.23-24.80); D-dimer 3.26 (1.31-7.05) and 13.89 (6.87-28.27); IL-6 2.61 (1.31-5.18) and 10.96 (5.88-20.43); ferritin 3.19 (1.66-6.11) and 9.13 (4.97-16.78); LDH 1.85 (0.87-3.97) and 10.51 (5.41-20.41); and NLR 3.34 (1.62-6.89) and 17.52 (9.03-34.00) (Table 8). We also calculated ORs (95% CI) for comparison of second and the third tertiles (Figure 3). Compared to the second tertile, the ORs were significantly greater in the third tertile for all the biomarkers-hsCRP 5.83 (3.72-9.14), D-dimer 4.26 (2.68-6.77), IL-6 4.22 (2.71-6.56), ferritin 2.86 (1.88-4.35), LDH 5.86 (3.30-9.548) as well as NLR 5.23 (3.45-7.92) (p<0.001).

**Table 8:** Univariate, age and sex-adjusted and multivariate adjusted association for in-hospital deaths in the second and third tertiles compared to the first and for third tertile compared to the second for various biomarkers.

|  | Unadjusted |  | Age-sex adjusted |  | Multivariate adjusted |  |  |
| --- | --- | --- | --- | --- | --- | --- | --- |
|  | Tertile 1 vs 2 | Tertile 1 vs 3 | Tertile 1 vs 2 | Tertile 1 vs 3 | Tertile 1 vs 2 | Tertile 1 vs 3 | Tertile 2 vs 3 |
| **hs-C reactive protein** | 2.34(1.17-4.70)* | 13.74(7.43-25.42)*** | 2.37(1.18-4.77)* | 14.30(7.70-26.55)*** | 2.29(1.14-4.60)* | 13.39(7.23-24.80)*** | 5.83(3.72-9.14)*** |
| **D-dimer** | 3.34(1.54-7.21)** | 14.34(7.06-29.13)*** | 3.20(1.48-6.92)** | 13.98(6.87-28.42)*** | 3.26(1.31-7.05)** | 13.89(6.87-28.27)*** | 4.26(2.68-6.77)*** |
| **Interleukin-6** | 2.64(1.33-5.25)** | 11.43(6.14-21.26)*** | 2.66(1.34-5.29)** | 11.41(6.13-21.24)*** | 2.61(1.31-5.18)** | 10.96(5.88-20.43)*** | 4.22(2.71-6.56)*** |
| **Ferritin** | 3.17(1.65-6.07)*** | 8.88(4.84-16.23)*** | 3.68(1.90-7.12)*** | 11.68(6.23-21.90)*** | 3.19(1.66-6.11)*** | 9.13(4.97-16.78)*** | 2.86(1.88-4.35)*** |
| **Lactic dehydrogenase** | 1.90(0.89-4.06) | 10.45(5.39-20.26)*** | 1.93(0.90-4.13) | 10.75(5.52-20.93)*** | 1.85(0.87-3.97) | 10.51(5.41-20.41)*** | 5.62(3.30-9.58)*** |
| **Neutrophils-Lymphocyte ratio** | 3.50(1.70-7.21)** | 18.32(9.45-35.50)*** | 3.47(1.68-7.15)** | 19.63(10.08-38.23)*** | 3.34(1.62-6.89)** | 17.52(9.03-34.0)*** | 5.23(3.45-7.91)*** |

**Figure 1:**
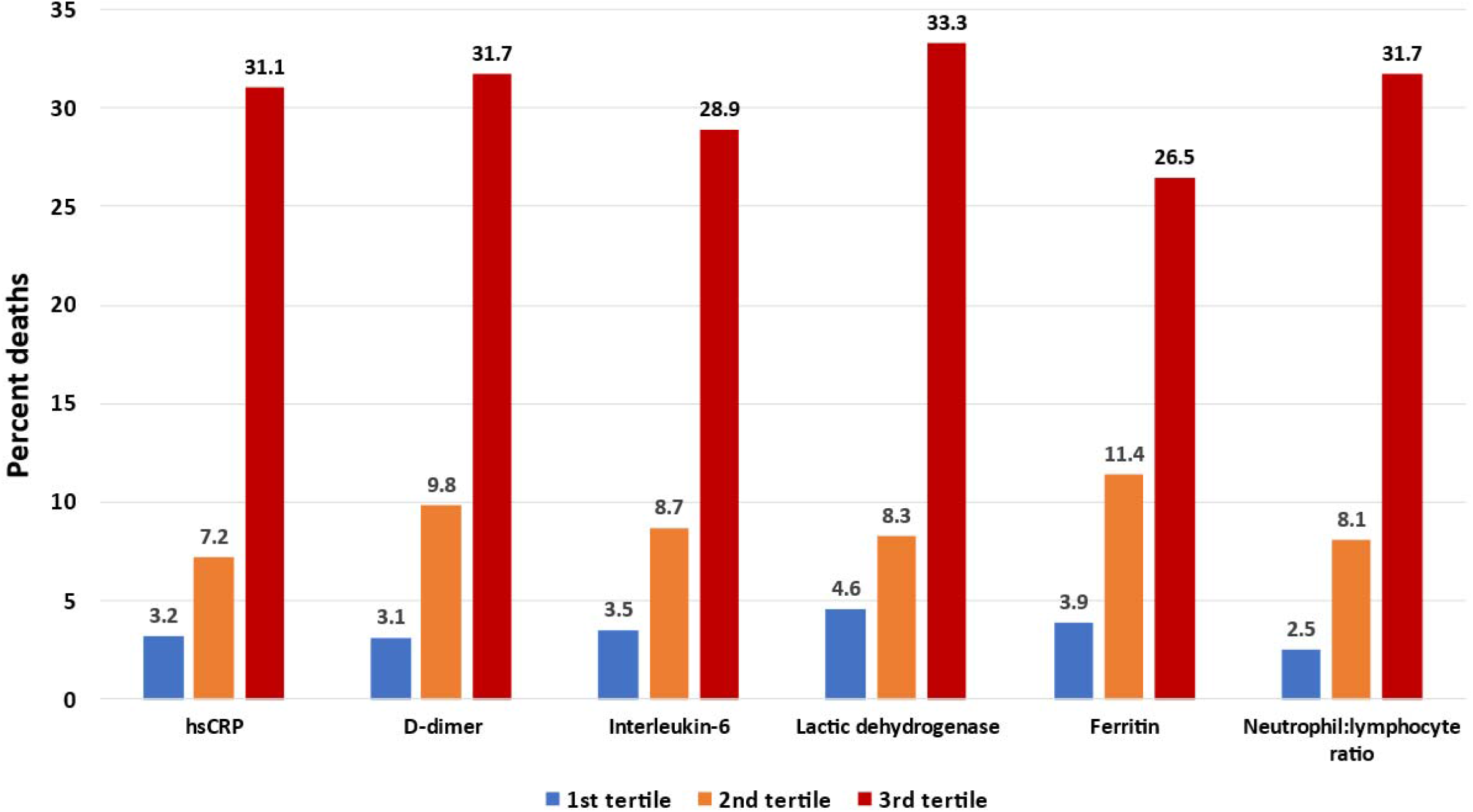
In-hospital deaths (%) in various biomarkers tertiles (hsCRP high sensitivity C-reactive protein)

**Figure 2:**
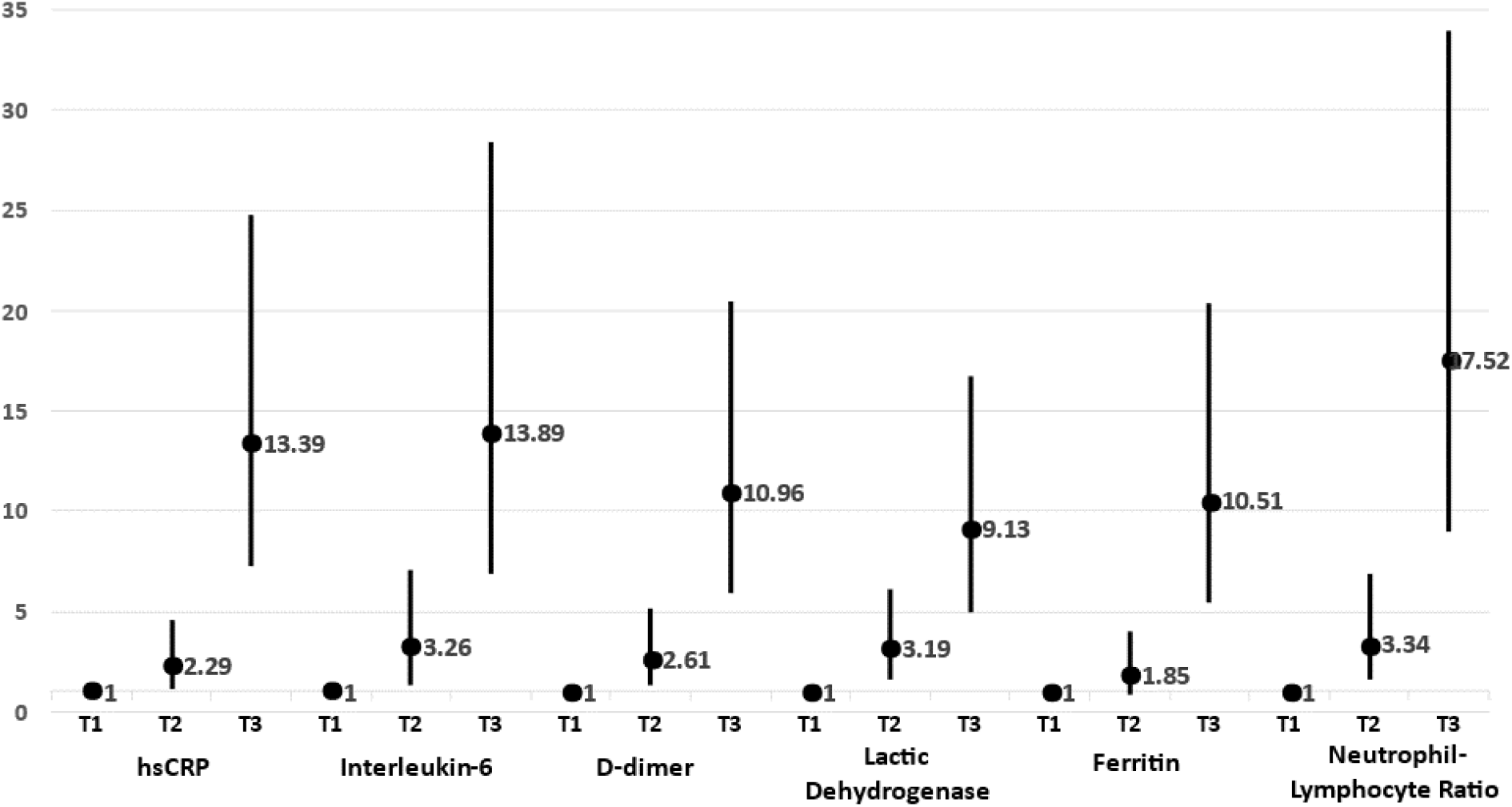
Multivariate (age, sex, risk factors, comorbidity) adjusted odds ratios and 95% confidence intervals for association of various biomarkers with in-hospital mortality. T1 (reference), T2 and T3 are the respective tertiles.

**Figure 3:**
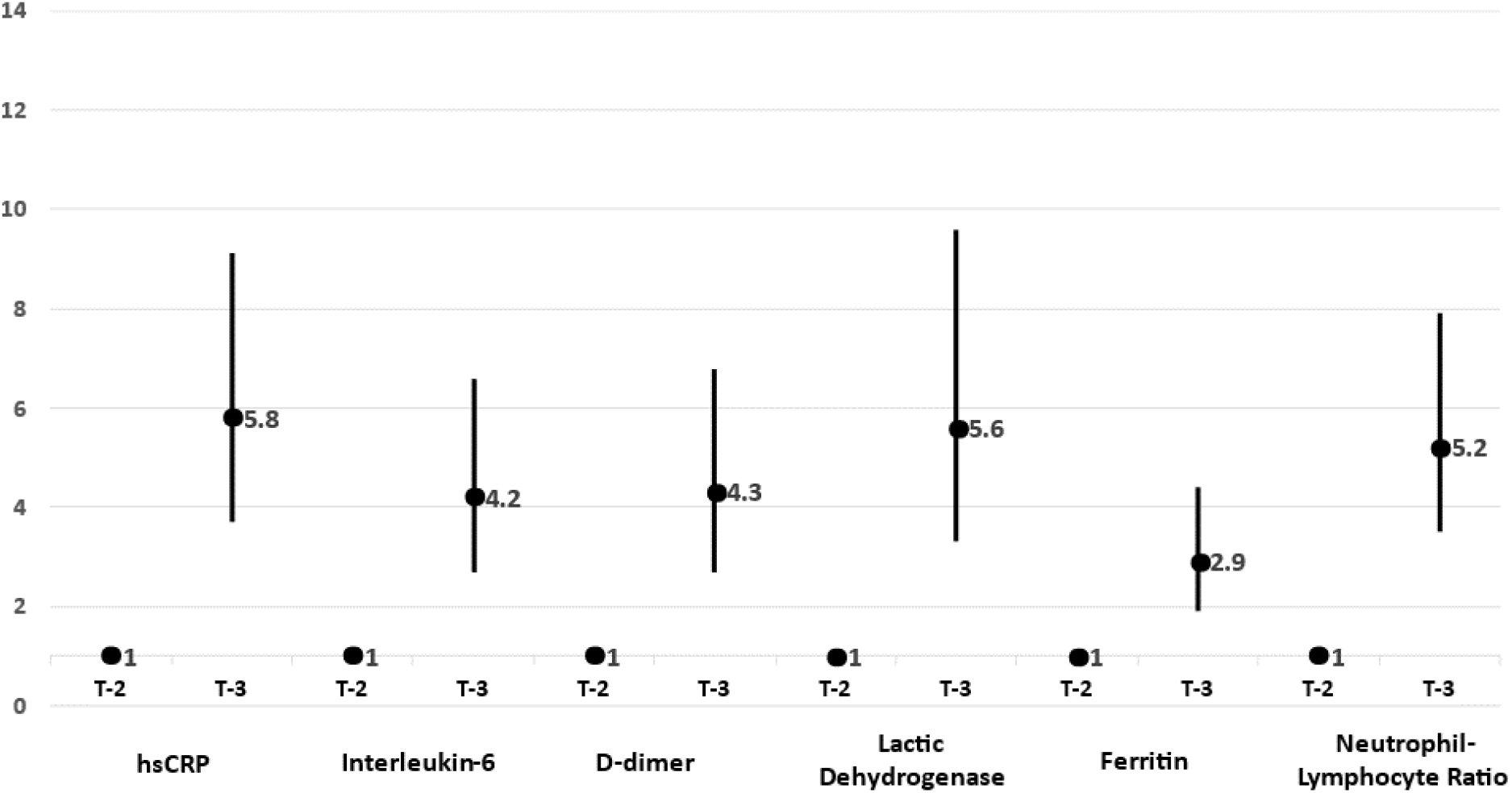
Multivariate (age, sex, risk factors, comorbidity) adjusted odds ratios and 95% confidence intervals for association of various biomarkers with in-hospital mortality on comparison of second tertile (T2) with the third (T3).

## DISCUSSION

This study shows that among consecutive patients hospitalized with Covid-19, increasing levels of biomarkers: hsCRP, D-dimer, interleukin-6, lactic dehydrogenase and ferritin as well as high neutrophil:lymphocyte ratio are associated with greater illness severity, oxygenation, non-invasive and invasive ventilation and exponentially greater in-hospital mortality. We also show that NLR, a simple, widely available and inexpensive investigation provides prognostic information that is similar to the more expensive biomarkers.

A meta-analysis that evaluated influence of biomarkers on poor outcomes (measured as oxygen saturation <90%, invasive mechanical ventilation, severe disease, ICU admission and mortality) among hospitalized patients with Covid-19 from December 2019 to August 2020 included 32 studies with 10491 patients.^9^ Biomarkers included were lymphocyte, platelets, D-dimer, LDH, CRP, AST, ALT, creatinine, procalcitonin and creatine kinase. This study reported a significant association between lymphopenia, thrombocytopenia and elevated levels of CRP, procalcitonin, LDH, D-dimer with COVID-19 severity with odds ratios varying from 3.33 (2.51-4.41) for lymphopenia, 4.37 (3.37-5.68) for elevated CRP and 5.48 (3.89-7.71) for LDH. We used mortality as the end-point and the results of our study are not directly comparable, however, the ORs are similar when we compared tertile 2 with tertile 3 in our study (Figure 3). Another meta-analysis reviewed impact of biomarkers of endothelial dysfunction (von Willebrand factor, tissue type plasminogen activator, soluble thrombomodulin, plasminogen activator inhibitor-1) and reported significant association with poor outcomes among 1187 patients from 17 studies.^23^ We did not evaluate these biomarkers and cannot directly compare our results, however, significance of D-dimer, another marker of coagulation and vascular dysfunction, in the present study suggests importance of coagulation markers in Covid-19 (Figure 2). A more recent review reported that biomarkers useful for risk prediction in Covid-19 include several proinflammatory cytokines, neuron-specific enolase, LDH, AST, neutrophil count, NLR, troponins, creatine kinase-MB, myoglobin, D-dimer, brain natriuretic peptide and N-terminal pro-hormone. Some of these biomarkers can be readily used to predict disease severity, hospitalization, ICU admission and mortality, while markers of metabolomic and proteomic analysis have not yet translated to clinical practice.^24^ The International Covid-19 Thrombosis Biomarkers Colloquim and international agencies recommend routine use of some of these biomarkers for prognostication.^8,22^ Our study shows a strong correlation among various biomarkers (Table 7) and suggests that clinical use of any one or two of these biomarkers may be enough for prognostication.

Our study also shows that a simple and inexpensive biomarker-NLR-is as predictive as other biomarkers. This is similar to previous studies,^9,24^ although the magnitude of risk is much higher as compared to the previous studies (Figure 2) but similar to previous studies when we compared the second and the third tertiles (Figure 3). Regolo et al,^25^ compared NLR with CRP in 411 elderly patients with Covid 19, 33% patients died during hospitalization. When divided into tertiles according to NLR values, it was observed that NLR was a better predictor of mortality than CRP with largest area under the curve (0.772) and high specificity (71.9%) and sensitivity (72.9%). Other studies, considerably smaller than ours, have also identified role of NLR in identification of severe disease.^26,27^ Reviews have concluded that this is an important marker,^7,8,23,28^ and the present study which is a larger than most of the previous studies, confirms this observation.

Our study has limitations and strengths, apart from those mentioned above. This is one of the larger studies that have evaluated multiple biomarkers in mild, moderate and severe Covid-19 cases and shows an exponential increase in deaths with rising levels of hsCRP, D-dimer, IL-6, LDH, ferritin and NLR and the strength of association is maintained after multivariate adjustment (Table 8). This is one of the larger studies from India, which has one of the highest deaths from Covid-19.^10,29,30^ Limitations include lack of assessment of other biomarkers, including some markers of coagulation and endothelial dysfunction mentioned above.^8,9,2324^ We also do not have long term data in these patients and cannot comment on importance of biomarkers in incidence of post-acute syndrome of Covid-19 (PASC). This is an important limitation as there are limited data that have addressed this question.^31,32^ Long term follow-up of this cohort can provide important answers.

In conclusion, the present study in hospitalized patients with Covid-19 shows that increasing levels of multiple biomarkers-hsCRP, D-dimer, IL-6, LDH, ferritin and NLR are associated with greater illness severity and significantly greater in-hospital mortality. The study also shows that, a universally available and inexpensive investigation, neutrophil:lymphocyte ratio, provide prognostic information similar to the more expensive biomarkers. Our study is important in the context of occurrence of multiple waves of Covid-19 in many developed and developing countries.^1-3^ The current omicron wave is due to its more pathogenic variants, BA.4, BA.5 and BA.2.75.^33,34^ We demonstrate that a simple investigation -neutrophil:lymphocyte ratio-can provide important prognostic information and can inexpensively triage patients presenting to primary care into low risk-who can be advised home-based care- and the intermediate and high risk groups to more intensive care settings.

## Data Availability

All the data produced in the present study are contained in the manuscript.

## Notes

### Competing Interest Statement

The authors have declared no competing interest.

### Funding Statement

The study did not receive any funding.

### Author Declarations

The protocol was approved by the institutional ethics committee of Eternal Heart Care Centre and Research Institute, Jaipur, India (Government of India registration, CDSCO No. ECR/615/Inst/RJ/2014/RR-20) before initiation of the study. Informed consent from all the patients or next-of-kin was obtained for anonymized data publication. The whole process was approved by the ethics committee.

## REFERENCES

1. Ritchie H, Mathieu E, Rodes-Guirao L. Appel C, Giattino C, Ortiz-Ospina E, et al. Coronavirus pandemic (Covid-19). Available at: https://ourworldindata.org/coronavirus. Accessed 21 Jun 2022.

2. Pagel C. The covid waves continue to come. BMJ 2022; 377:o1504.

3. Young M, Crook H, Scott J, Edison P. Covid-19: virology, variants and vaccines. BMJ Med. 2022; 1. doi: 10.1136/bmjmed-2021-000040.

4. Mohsin M, Mahmud S. Omicron SARS-CoV-2 variant of concern: a review on its transmissibility, immune evasion, reinfection and severity. Medicine (Baltimore). 2022; 101:e29165.

5. Guo Y, Han J, Zhang Y, He J, Yu W, Zhang X, et al. SARS-CoV-2 omicron variant: epidemiological features, biological characteristics, and clinical significance. Front Immunol. 2022; 13:877101/

6. Duong BV, Larpruenrudee P, Fang T, Hossain SI, Saha SC, Gu Y, et al. Is the SARS CoV-2 omicron deadlier and more transmissible than delta variant? Int J Environ Res Public Health. 2022; 19:4586.

7. Cao B, Jing X, Liu Y, Wen R, Wang C. Comparison of laboratory parameters in mild vs severe cases and died vs survived patients with Covid-19: systematic review and meta-analysis. J Thorac Dis. 2022; 14:1478–87.

8. Gorog DA, Storey RF, Gurbel PA, Tantry US, Berger JS, Chan MY, et al. Current and novel biomarkers of thrombotic risk in Covid-19: a consensus statement from the International Covid-19 Thrombosis Biomarkers Colloquium. Nature Rev Cardiol. 2022; 19:475–495.

9. Malik P, Patel U, Mehta D, Patel N, Kelkar R, Akrmah M, et al. Biomarkers and outcomes of Covid-19 hospitalizations: systematic review and meta-analysis. BMJ Evid Based Med. 2021; 26:107–118.

10. World Health Organization. The true death toll of Covid-19: estimating global excess mortality. Available at https://www.who.int/data/stories/the-true-death-toll-of-covid-19-estimating-global-excess-mortality. Accessed 21 Jun 2022.

11. Mammen JJ, Kumar S, Thomas L, Kumar G, Zachariah A, Jeyaseelan L, et al. Factors associated with mortality among moderate and severe patients with COVID-19 in India: a secondary analysis of a randomised controlled trial. BMJ Open. 2021;11:e050571.

12. Gupta D, Jain A, Chauhan M, Dewan S, Inflammatory markers as early predictors of disease severity in Covid-19 patients admitted to intensive care units: a retrospective observational analysis. Indian J Crit Care Med. 2022; 26:482–486.

13. Bohra GK, Bhatia PK, Khichar S, Garg MK, Sharma P. Association of inflammatory markers with Covid-19 outcome among hospitalised adult patients. J Assoc Physicians India. 2022; 70(4):11–12.

14. Bhandari S, Rankawat G, Mathur S, Kumar A, Sahlot R, Jain A. Circulatory cytokines as a predictor of disease severity in Covid-19: a study from western India. J Assoc Physicians India. 2022; 70(5):11–12.

15. Zemlin AE, Allwood B, Erasmus RT, Matsha TE, Chapanduka ZC, Jalavu TP, et al. Prognostic value of biochemical parameters among severe Covid-19 patients admitted to an intensive care unit of a tertiary hospital in South Africa. IJID Reg. 2022; 2:191–197.

16. Ansari N, Jahangiri M, Shirbandi K, Ebrahimi M, Rahim F. The association between different predictive biomarkers and mortality of Covid-19. Bull Natl Res Cent. 2022; 46:158.

17. Alle S, Kanakan A, Siddiqui S, Garg A, Karthikeyan A, Mehta P, et al. Covid-19 risk stratification and mortality prediction in hospitalised Indian patients: harnessing clinical data for public health benefits. PLoS One. 2022; 17:e0264785.

18. Khedar RS, Mittal K, Ambaliya HC, Mathur A, Gupta JB, Sharma KK, et al. Greater Covid-19 severity and mortality in hospitalized patients in the second (delta) wave compared to the first wave of epidemic: single centre prospective study in India. medRxiv preprints. 2021; doi: https://doi.org/10.1101/2021.09.03.21263091.

19. Sharma AK, Gupta R, Baig VN, Singh TV, Chakraborty S, Sunda JP, et al. Educational status and COVID-19 related outcomes in India: hospital based cross-sectional study. BMJ Open. 2022; 10:e055403.

20. Sharma S, Sharma AK, Dalela G, Dhakar P, Baig VN, Singh TV, et al. Association of SARS CoV-2 cycle threshold (Ct) with clinical coutcomes: hospital-based study. J Assoc Physicians India. 2021; 69(7):20–24.

21. Government of India, Ministry of Health. Clinical management protocol for Covid-19 in adults. 24<sup>th</sup> May 2021. Available at: https://www.mohfw.gov.in/pdf/UpdatedDetailedClinicalManagementProtocolforCOVID19adultsdated24052021.pdf. Accessed 7 July 2022.

22. Centers for Disease Control and Prevention. Clinical care guidance. Available at: https://www.cdc.gov/coronavirus/2019-ncov/hcp/clinical-care/clinical-considerations-index.html. Accessed 7 July 2022.

23. Andrianto A, Al-Farabi MJ, Nugraha RA, Marsudi BA, Azmi Y. Biomarkers of endothelial dysfunction and outcomes in Covid-19 patients: a systematic review and meta-analysis. Microvasc Res. 2021; 138:104224.

24. Battaglini D, Lopes-Pacheo M, Castro-Faria-Neto HC, Pelosi P, Rocco PRM. Laboratory biomarkers for diagnosis and prognosis in Covid-19. Front Immunol. 2022; 13:857573.

25. Regolo M, Vaccaro M, Stancanelli B, Colaci M, Natoli G, et al. Neutrophil-to-lymphocyte ratio (NLR) is a promising predictor of mortality and admission to intensive cafre unit of Covid-19 patients. J Clin Med. 2022; 11:2235.

26. Prakash Rao VV. Utility of neutrophil-lymphocyte ratio (NLR) as an indicator of disease severity and prognostic marker among patients with Covid-19 infection ina tertiary care centre in Bangalore-a retrospective study. J Assoc Physicians India. 2022; 70(4):11–12.

27. Abdelhady SA, Rageh F, Ahmed SS, Al-Touny SA, Riad E, Elhoseeny MM, et al. Neutrophil to lymphocyte ratio and other inflammatory marketrs as adverse outcome predictor in hospitalized Covid-19 patients. Egypt J Immunol. 2022; 29:57–67.

28. Buonacera A, Stancanelli B, Colaci M, Malatino L. Neutrophil to lymphocyte ratio: an emerging marker of the relationships between the immune system and diseases. Inj J Mol Sci. 2022; 23:3636.

29. Jha P, Deshmukh Y, Tumbe C, Suraweera W, Bhowmick A, Sharma S, et al. COVID mortality in India: National survey data and health facility deaths. Science. 2022; 375:667–671.

30. Covid-19 Cumulative Infection Collaborators. Estimating global, regional and national daily and cumulative infections with SARS-CoV-2 through Nov 14, 2021: a statistical analysis. Lancet. 2022; EPub. DOI: https://doi.org/10.1016/S0140-6736(22)00484-6.

31. Nalbandian A, Sehgal K, Gupta A, Madhavan MV, McGroder C, Stevens JS, et al. Post-acute Covid syndrome. Nature Med. 2021; 27:601–615.

32. Hope AA, Evering TH. Post-acute sequelae of severe acute respiratory syndrome coronavirus-2 infection. Infect Dis Clin North Am. 2022; 36:379–395.

33. Topol E. The BA.5 story: the takeover by this omicron sub-variant is not pretty. Available at: https://erictopol.substack.com/p/the-ba5-story. Accessed 7 July 2022.

34. Prater E. New omicron subvariant centaurus could be the most immune-evasive yet, expert warns. Available at: https://fortune.com/2022/07/06/new-omicron-subvariant-centaurus-ba-2-75-world-health-organization-immune-evasive-expert/. Accessed 7 July 2022.

